# Infectiousness in omicron variant strain and bA.2 variant in Japan

**DOI:** 10.1101/2021.06.20.21259209

**Authors:** Junko Kurita, Tamie Sugawara, Yasushi Ohkusa

## Abstract

**Background:** Omicron variant strain dominated since the beginning of 2022. Its infectivity was supposes to be higher than Delta variant strain or strains in past.

**Object:** We estimated prevalence of omicron variant strain, particularly bA.2 variant and COVID-19 vaccine effectiveness of the third dose in Japan as well as controlling for waning of second dose of vaccine, other mutated strains, the Olympic Games, and countermeasures.

**Method:** The effective reproduction number R(*t*) was regressed on shares of omicron variant strain and bA.2 and vaccine coverage of the third dose, as well as along with data of temperature, humidity, mobility, share of the other mutated strains, and an Olympic Games and countermeasures. The study period was February, 2020 through February 21, 2022, as of March 15, 2022.

**Results:** Estimation results indicated that waning of the second dose vaccine e with 150 days prior was the most appropriate specification. Moreover, bA.2 of omicron variant strain has higher infectively than other variant strain or traditional strain.

**Discussion:** Because of data limitation since emerging bA.2, the estimated its infectively will change over time.

## 1. Introduction

Omicron variant strain dominated since the beginning of 2022 in Japan as well as rest of the world. Though some researched supposed higher infectivity than Delta variant strain or strains in past [1,2], it was much concern of public health and general in the real world, especially in the community as well as its pathogenicity. Moreover, since February 2020, bA.2 strain mutated from omicron variant strain [3-5].

Before delta variant strain emerging, wide coverage of COVID-19 vaccination has altered outbreak situations in European countries and in the US. Unfortunately, vaccination in Japan started only in February, 2021 using BNT162b2 mRNA (Pfizer Inc., BioNTech) and mRNA-1273 (Moderna, Inc.) vaccines: among the latest of starting dates of vaccination programs in economically developed countries. Later, ChAdOx1 adenoviral vector (Oxford, AstraZeneca) also became available. By the end of November 2021, the rate of completion for second dose vaccine administration had reached almost 80% in Japan (Figure 1) [6,7]. The next challenge posed by vaccine issues in Japan might be waning of vaccine effectiveness.

**Figure 1:**
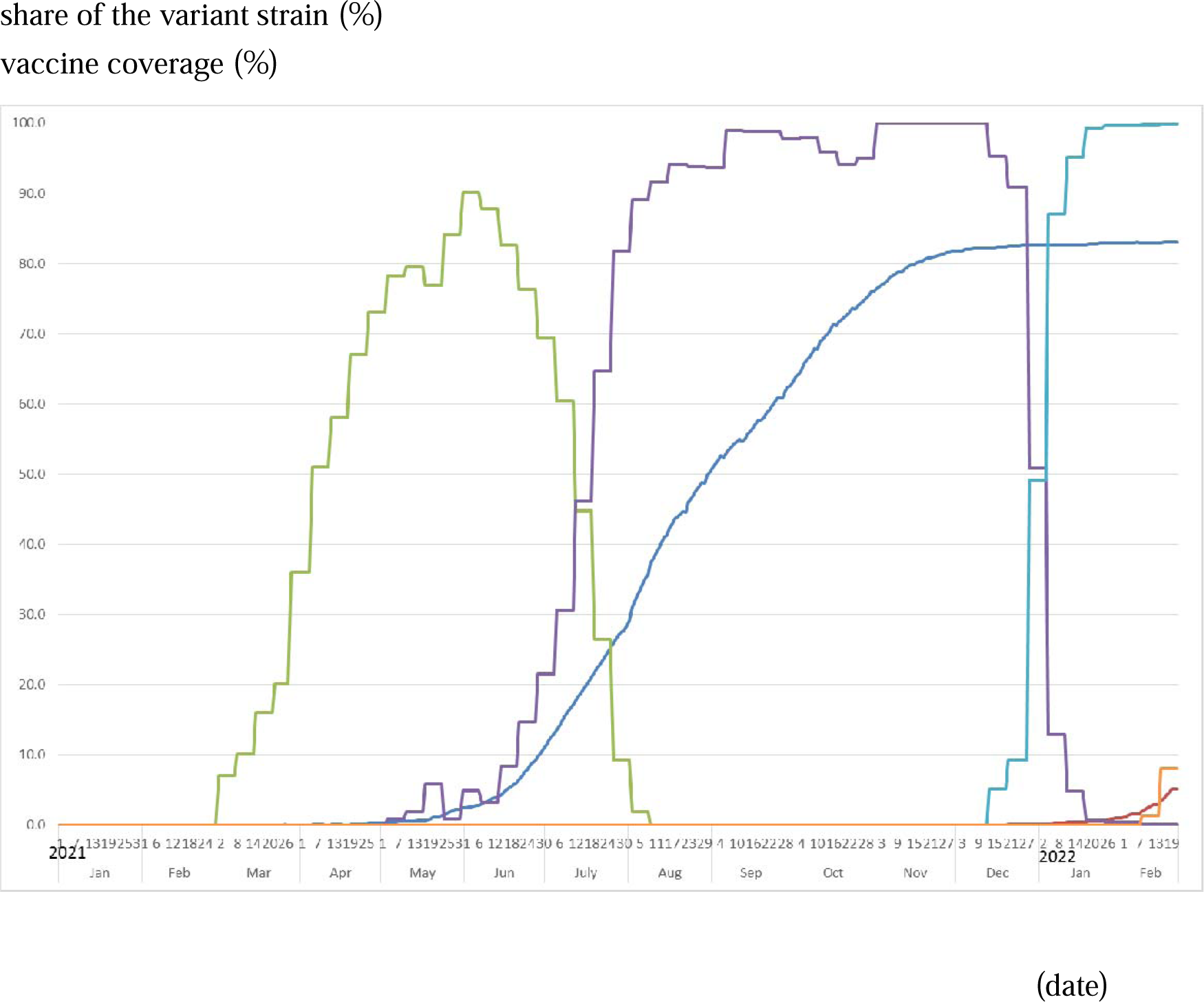
Vaccine coverage and shares of alpha and delta variant strains in 2021 until February 21, 2022. Note: The green line represents shares of the alpha variant strain, the purple line represents shares of the delta variant strain, and light blue line indicates the omicron variant strain including bA.2 in Tokyo. Red line indicates share of only bA.2 variant strain. Blue line denote completed vaccine coverage as defined by the second dose with a 14-day delay. Red line denote vaccine coverage defined by the third dose. Because the daily vaccine coverage was not reported on weekends or national holidays, data of vaccine coverage are missing for these days. Moreover, there were adjustments for double counting for the number of vaccine recipients. Therefore, the vaccine coverage sometimes slightly decrease from before.

In fact, waning vaccine effectiveness has been reported [8,9]. One study revealed that the log of IgG antibody titer decreased by a factor of 18.3 when measured six months after second-dose vaccination. Another study revealed vaccine effectiveness as 77.5% at one month after the second vaccination, but it had decreased to about 20% when measured 5–7 months later. In the real world, vaccines of several types have been used. Moreover, vaccinated persons might change their behaviors. Therefore, this study assessed vaccine effectiveness and its waning capabilities against infectively in the real world, particularly in Japan.

By the time vaccinations started in Japan, the alpha variant strain had emerged and had expanded to dominate the recorded infections. Subsequently, a new mutant alpha variant strain appeared in May. Based mainly on data reported by the UK, its infectively and pathogenicity were estimated as 35–90% higher than those of the original strain circulating before the emerging variant strain [10-13]. Therefore, we consider the prevalence of these mutated strains together when evaluating vaccine effects.

The Olympic Games and Paralympic Games of 2020 began on July 23, 2021. A subject of great concern for COVID-19 outbreak effects in Japan was whether audiences would be allowed to attend game events, or not. As part of this controversy, some experts asserted that the Games should be abandoned because they would expand the outbreak explosively [14].

As a result, the game events were held with no live audience. Under the state of emergency declared in Tokyo, effects of the 2020 Tokyo Games must be included to evaluate vaccine effectiveness.

As countermeasures against the COVID-19 outbreak in Japan, school closure and voluntary event cancellation were adopted from February 27, 2020 through the end of March. Large commercial events were cancelled. Subsequently, a state of emergency was declared for April 7 through 25 May, stipulating voluntary restrictions against leaving home. Consumer businesses such as retail shops and restaurants were shuttered. During this period, the first peak of infection was reached on April 3. Infections subsequently decreased through July 29. The so-called “Go To Travel Campaign” (GTTC) was launched on July 22 as a 50% subsidized travel program aimed at supporting sightseeing and tourism businesses with government-issued coupons for use in shopping at tourist destinations. It was expected that the campaign might expand the outbreak. Thereafter, GTTC continued to the end of December, by which time a third wave of infection had emerged. The third wave in December, which was larger than either of the preceding two waves, reached its highest peak at the end of December. Therefore, GTTC was inferred as the main reason underlying the third wave [15].

To suppress that third wave of infection, a second state of emergency was declared from January 8, 2021 through March 15, 2021. However, a fourth wave emerged at the end of February, probably because of the spread of variant strains. To support hosting of the Olympics and Paralympics games in Tokyo in July, a third state of emergency was declared on April 25, 2021. It had ceased on June 20, 2021 in Tokyo. Nevertheless, the outbreak commenced again before the Tokyo Games 2020 started. Therefore, a fourth state of emergency was declared on July 13, 2021. It continued thereafter until the Tokyo Games 2020 had closed.

Although results have been mixed, some findings from earlier studies suggest that COVID-19 is associated with climate conditions [16–19]. If that were true for Japan, then GTTC might not have been the main factor contributing to the third wave. In fact, mobility was inferred as the main cause of the outbreak dynamics for the first wave in Japan [20] and throughout the world [21–24].

The object of this study was to estimate waning of vaccine effectiveness against SARS-CoV-2 infectively for the outbreak in Japan as a result of the vaccine effectiveness itself, the mutated strain, the Olympic Games, countermeasures, and other factors that might affect infectively.

## 2. Methods

This study examined the numbers of symptomatic patients reported by the Ministry of Health, Labour and Welfare (MHLW) for February 1, 2020 – February 21, 2022 published [25]as of March 15, 2022. Some patients were excluded from data for Japan: patients presumed to be persons infected abroad or infected as Diamond Princess passengers. Those patients were presumed not to represent community-acquired infection in Japan. For some symptomatic patients with unknown onset dates, we estimated the onset dates from an empirical distribution with duration extending from onset to the report date among patients for whom the onset date had been reported.

Onset dates among patients who did not report this information and a reporting delay were adjusted using the same procedures as those used for earlier studies [26,27]. As described hereinafter, we estimated the onset dates of patients for whom onset dates had not been reported. Letting *f*(*k*) represent this empirical distribution of the incubation period and letting *N*_*t*_ denote the number of patients for whom onset dates were not published and available at date *t*, then the number of patients for whom the onset date was known is *t*-1. The number of patients with onset date *t*-1 for whom onset dates were not available was estimated as *f*(1)*N*_*t*_. Similarly, patients with onset date *t*-2 and for whom onset dates were not available were estimated as *f*(2)*N*_*t*_. Therefore, the total number of patients for whom the onset date was not available, given an onset date of *s*, was estimated as Σ_*k*=1_*f*(*k*)*N*_*s*_+*k* for the long duration extending from *s*.

Moreover, the reporting delay for published data from MHLW might be considerable. In other words, if *s*+*k* is larger than in the current period *t*, then *s*+*k* represents the future for period *t*. For that reason, *N*_*s*+*k*_ is not observable. Such a reporting delay engenders underestimation of the number of patients. For that reason, it must be adjusted as Σ_*k*=1_^*t*-*s*^*f*(*k*)*N*_*s*_*+k* /Σ_*k*=1_^*t*-*s*^*f*(*k*). Similarly, patients for whom the onset dates were available are expected to be affected by the reporting delay. Therefore, we have *M*_*s*_|_*t*_ /Σ_*k*=1_^*t*-*s*^*f*(*k*), where *M*_*s*_|_*t*_ represents the reported number of patients for whom onset dates were period *s* as of the current period *t*. We defined R(*t*) as the number of infected patients on day *t* divided by the number of patients who were presumed to be infectious. The number of infected patients was calculated from the epidemic curve by the onset date using an empirical distribution of the incubation period, which is Σ_*k*=*1*_*f*(*k*)*E*_*t+k*_, where *E*_*t*_ denotes the number of patients for whom the onset date was period *t*. The distribution of infectively in symptomatic and asymptomatic cases *g*(*k*) was assumed to be 30% on the onset day, 20% on the following day, and 10% for the subsequent five days [28]. Then the number of infectious patients was Σ_*k*=1_*g*(*k*)*E*_*t*-*k*_. Therefore, R(*t*) was defined as Σ_*k*=1_*f*(*k*)*E*_*t+k*_/Σ_*k*=1_*g*(*k*)*E*_*t*-*k*_.

Data indicating the shares of mutated variants among all cases were published by the Tokyo Metropolitan Government. Unfortunately, detailed information about mutated strains has not been published for the entirety of Japan. We used four measures for the mutant strain shares in Tokyo, Japan: alpha, delta, omicron and bA.2 variant strains [29].

We use average temperature and relative humidity data for Tokyo during the day as climate data because national average data are not available. We obtained data from the Japan Meteorological Agency (https://www.data.jma.go.jp/gmd/risk/obsdl/index.php). Additionally, we identified several remarkable countermeasures in Japan: four state-of-emergency declarations, a travel campaign, and school closure and voluntary event cancellation (SCVEC). The latter, SCVEC, extended from February 27 through March in 2020: this countermeasure required school closure and cancellation of voluntary events, and even cancellation of private meetings. The first state of emergency was declared on April 7, 2020. It ceased at the end of May. It required school closures, shutting down of some businesses, and voluntary restriction against going out. To subsidize travel and shopping at tourist destinations, the “Go To Travel Campagn (GTTC)” started on July 22, 2020. It was halted temporarily at the end of December.

The second state of emergency was declared on January 7, 2021 for the 11 most-affected prefectures. This countermeasure required restaurant closure at 8:00 p.m., with voluntary restrictions against going out, but it did not require school closure. It continued until March 21, 2021. The third state of emergency was declared on April 25, 2021 for four prefectures: Tokyo, Osaka, Hyogo, and Kyoto. Later, the application areas were extended gradually. They never covered the entirety of Japan.

To clarify associations among R(*t*) and current and the past vaccine coverage in addition to the mutant strains, climate, mobility, the Olympic Games, and countermeasures, we used ordinary least squares regression to regress the daily R(*t*) on daily current vaccine coverage and daily past vaccine coverage as well as dummy variables for the Games, weekly shares of alpha and delta variant strains, daily climate, mobility, and dummy variables for countermeasures. Temperatures were measured in degrees Celsius, with humidity and mobility as percentages in regression, not as standardized. Variables found to be not significant were excluded from explanatory variables. Then the equation was estimated again.

We define vaccine coverage as the completion rate of the second dose without delay. If a vaccine perfectly protects the recipient from infection, then the estimated coefficient of vaccine coverage would be 0.01 if one assumes an average of R(*t*) with no vaccination in the study period. That would indicate that vaccine coverage increased by one percentage point could be expected to reduce R(*t*) by one percentage point. If the estimated coefficient of vaccine coverage were smaller than -0.01, then it might reflect imperfect personal prevention. Conversely, if the estimated coefficients of vaccine coverage were smaller than -0.01, then herd immunity can be inferred to have contributed to prevention of infection among non-recipients.

Waning of vaccine effectiveness was measured by the estimated coefficient of vaccine coverage in the past. Particularly, we examined every 30 days prior until 180 days prior. We expected the estimated coefficient to be positive if waning was occurring. If its estimated coefficient was positive but smaller than the absolute value of the estimated coefficient of current vaccine coverage, then waning was presumed to be partially occurring. Vaccination was presumed to be effective even if a part of effectiveness was waning. If the estimated coefficient of vaccine coverage in the past was positive and almost equal to the absolute value of the estimated coefficient of current vaccine coverage, then waning was presumed to be complete. We might not expect vaccine effectiveness until that time. Conversely, if the estimated coefficient of vaccine coverage in the past was positive and larger than the absolute value of the estimated coefficient of current vaccine coverage, then the vaccine might raise infectively eventually. We adopted 5% as the level at which we inferred significance of the results.

## 3. Results

### 3.1 Data

Figure 1 depicts vaccine coverage for the first dose with a 12-day delay and depicts the second dose as scatter diagrams. It also shows the shares of alpha and delta variant strains as bars. These are increasing almost monotonically during the period. Adjustments were made for double counting for the number of vaccine recipients. Therefore, the vaccine coverage was sometimes less than it was earlier. Figure 2 depicts R(*t*) during the study period.

**Figure 2:**
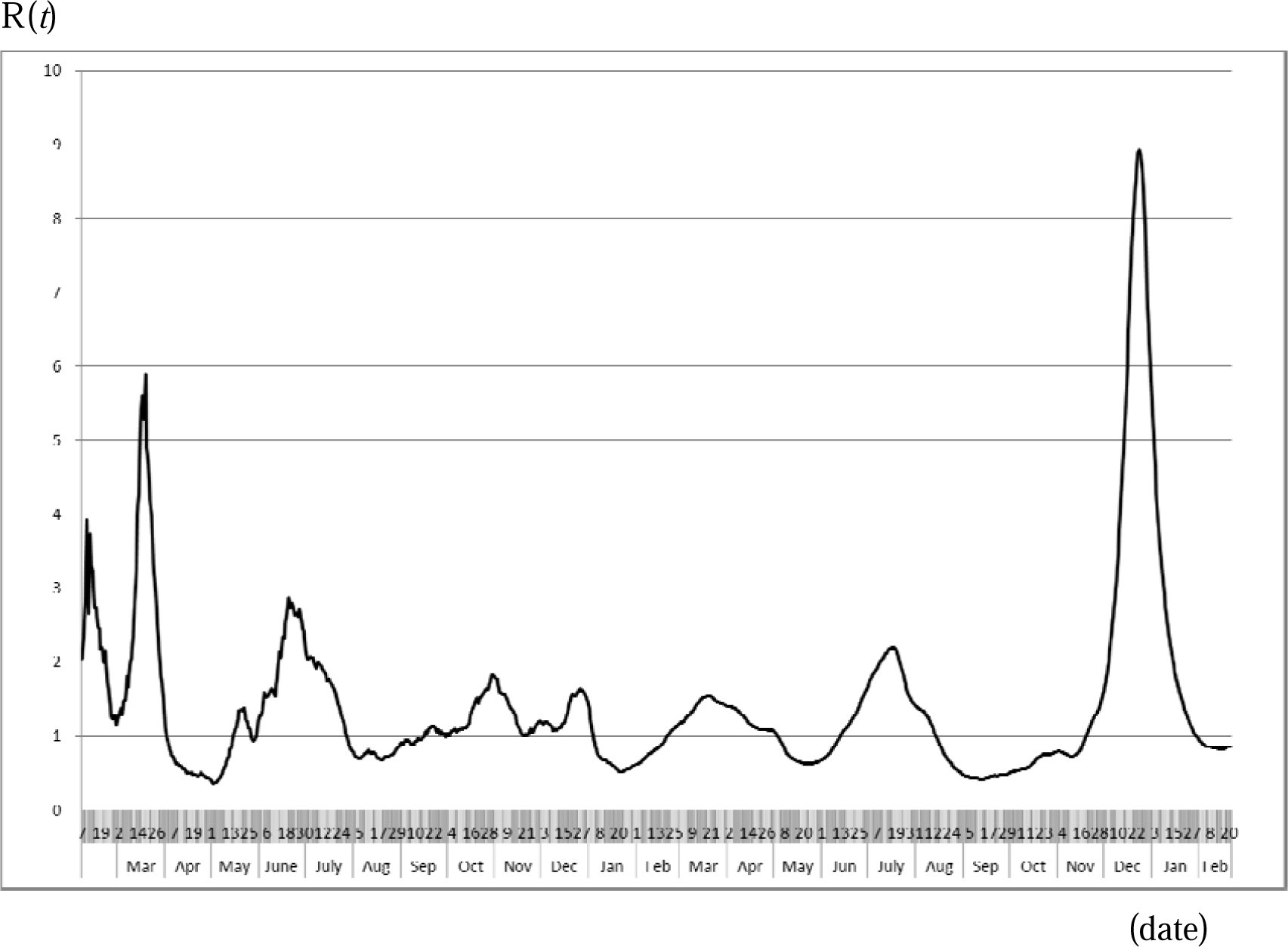
Effective reproduction number from February, 2020 through February 21, 2022. Note: The line represents the effective reproduction number in Japan from February, 2020 through February 21, 2022, as of March 15,2020. Calculation procedures are explained in the main text.

Figure 3 presents an empirical distribution of the duration of onset to reporting in Japan. The maximum delay was 31 days. Figure 4 presents an empirical distribution of incubation periods among 91 cases for which the exposed date and onset date were published by MHLW in Japan. The mode was six days; the average was 6.6 days.

**Figure 3:**
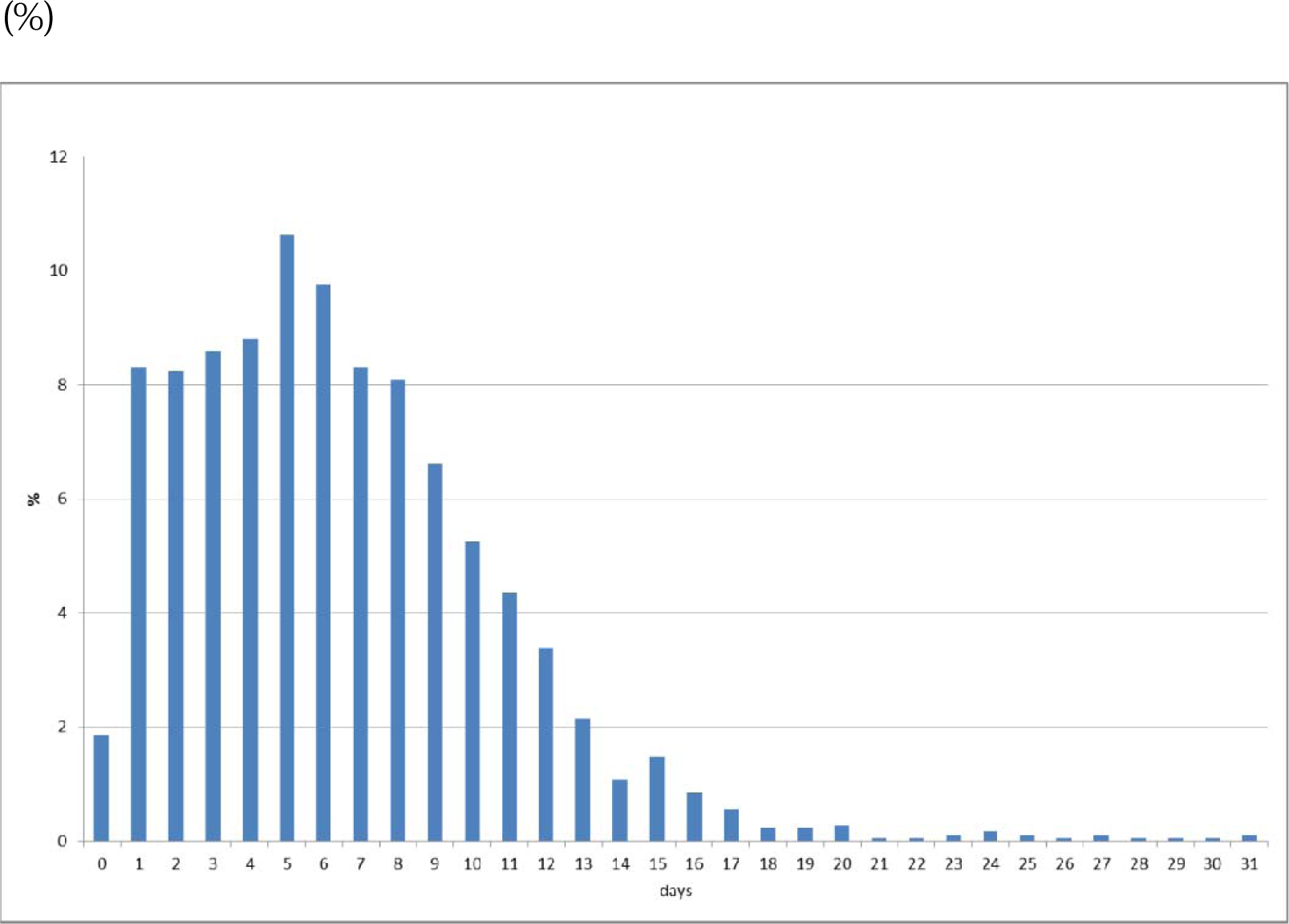
Empirical distribution of duration from onset to report by MHLW, Japan. Note: Bars represent the probability of duration from onset to report based on 657 patients in Japan for whom the onset date was available. Data were obtained from MHLW, Japan.

**Figure 4:**
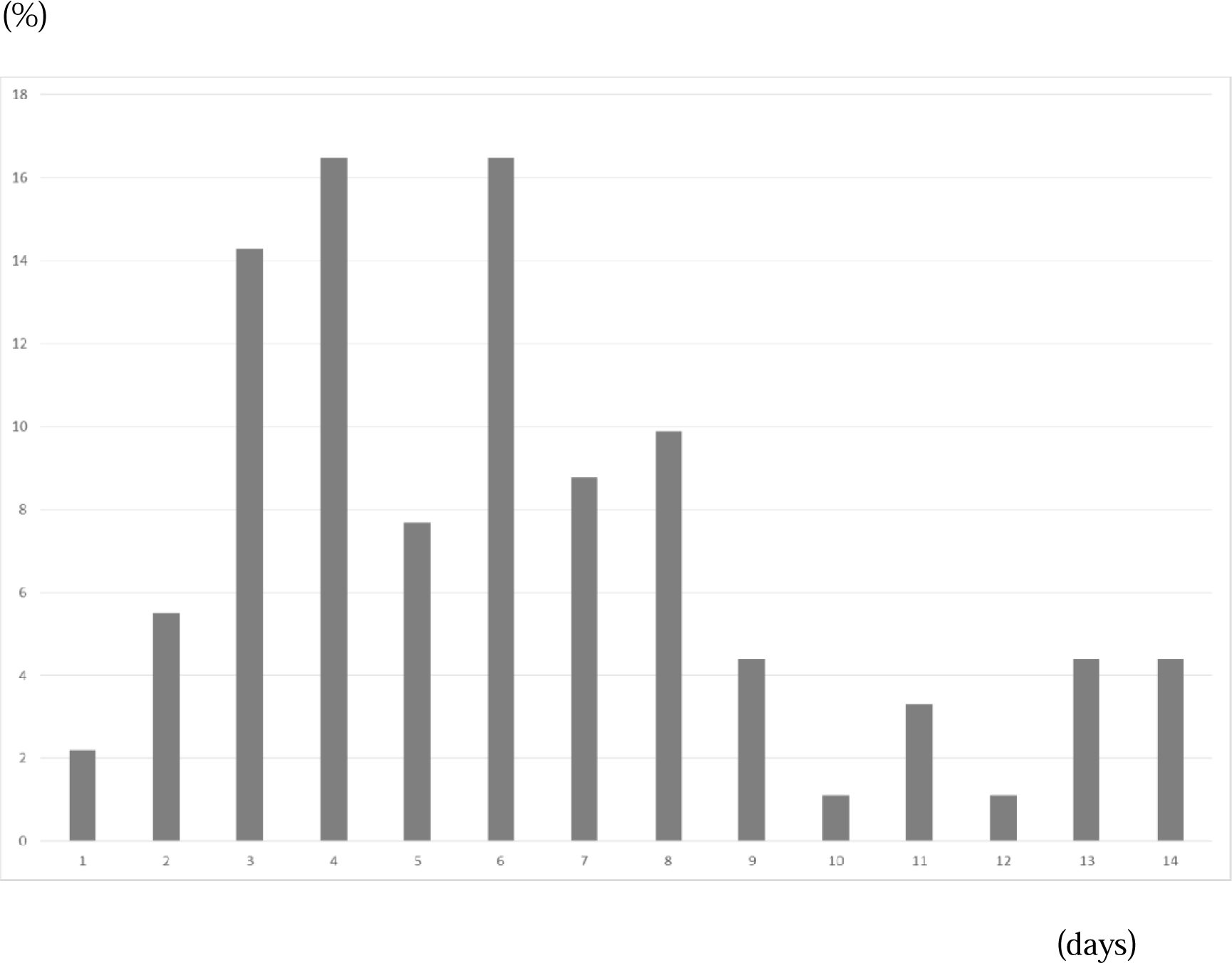
Empirical distribution of the incubation period published by MHLW, Japan. Notes: Bars show the distribution of incubation periods for 91 cases for which the exposure date and onset date were published by MHLW, Japan. Patients for whom incubation was longer than 14 days are included in the bar shown for day 14.

Table 1 presents estimation results. Based on the estimated adjusted *R*^*2*^, we selected the specification with 150 days lag of waning of the second dose vaccination. In this specification, mobility, SCVEC, vaccine coverage of the second dose with lag, and share of delta or omicron bA.2 variant strain were positively significant. Conversely, 1^st^ to 3rd state of emergency, GTTC, vaccine coverage of the second and third dose, share of alpha or omicron including bA,2 variant strain was negatively significant. Climate conditions, 4th State of emergency, and Olympic Games were not significant. Especially, bA.2 showed very high infectively even though infectively of omicron was lower than traditional strain before alpha variant strain.

**Table 1:**
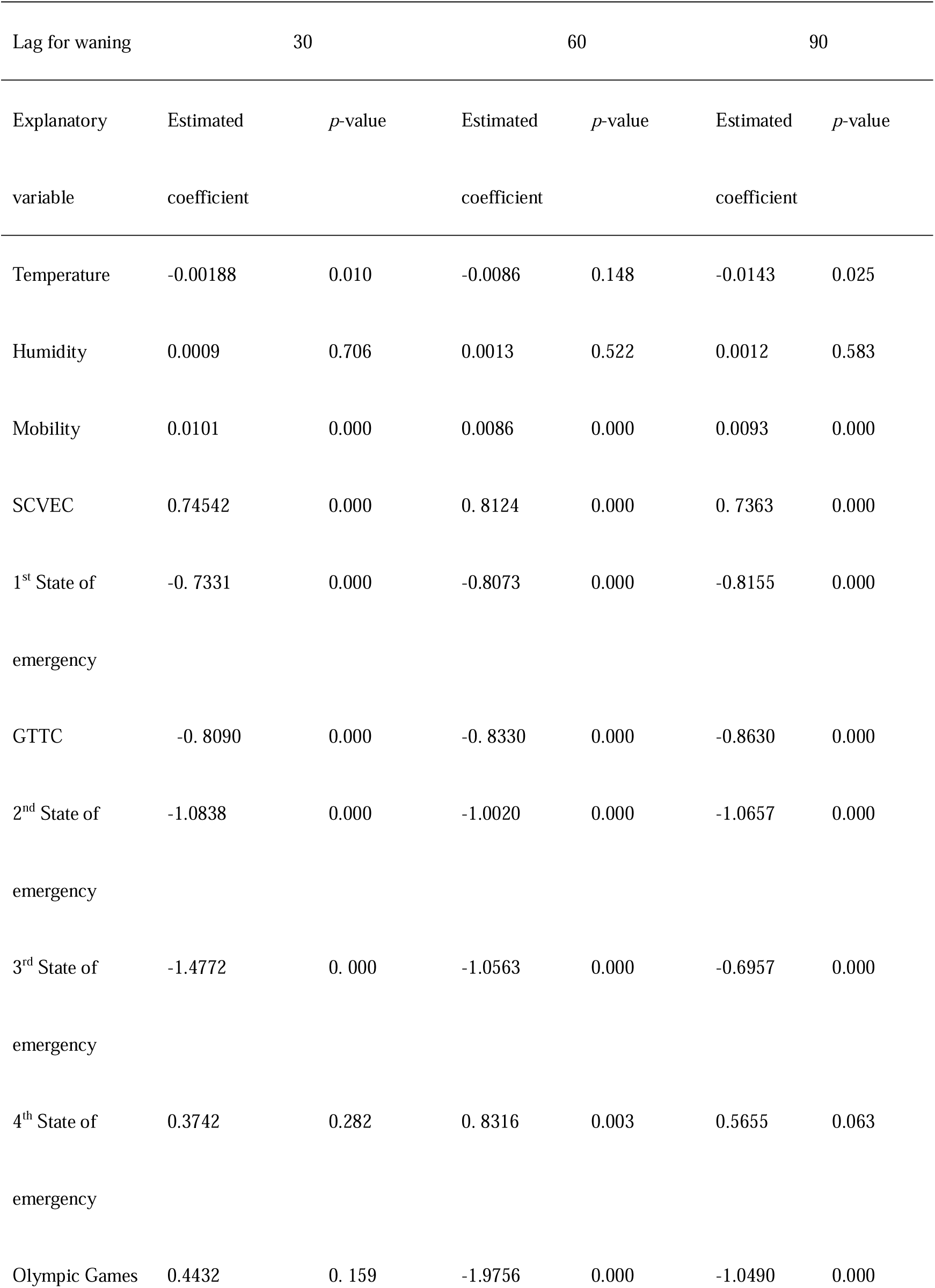
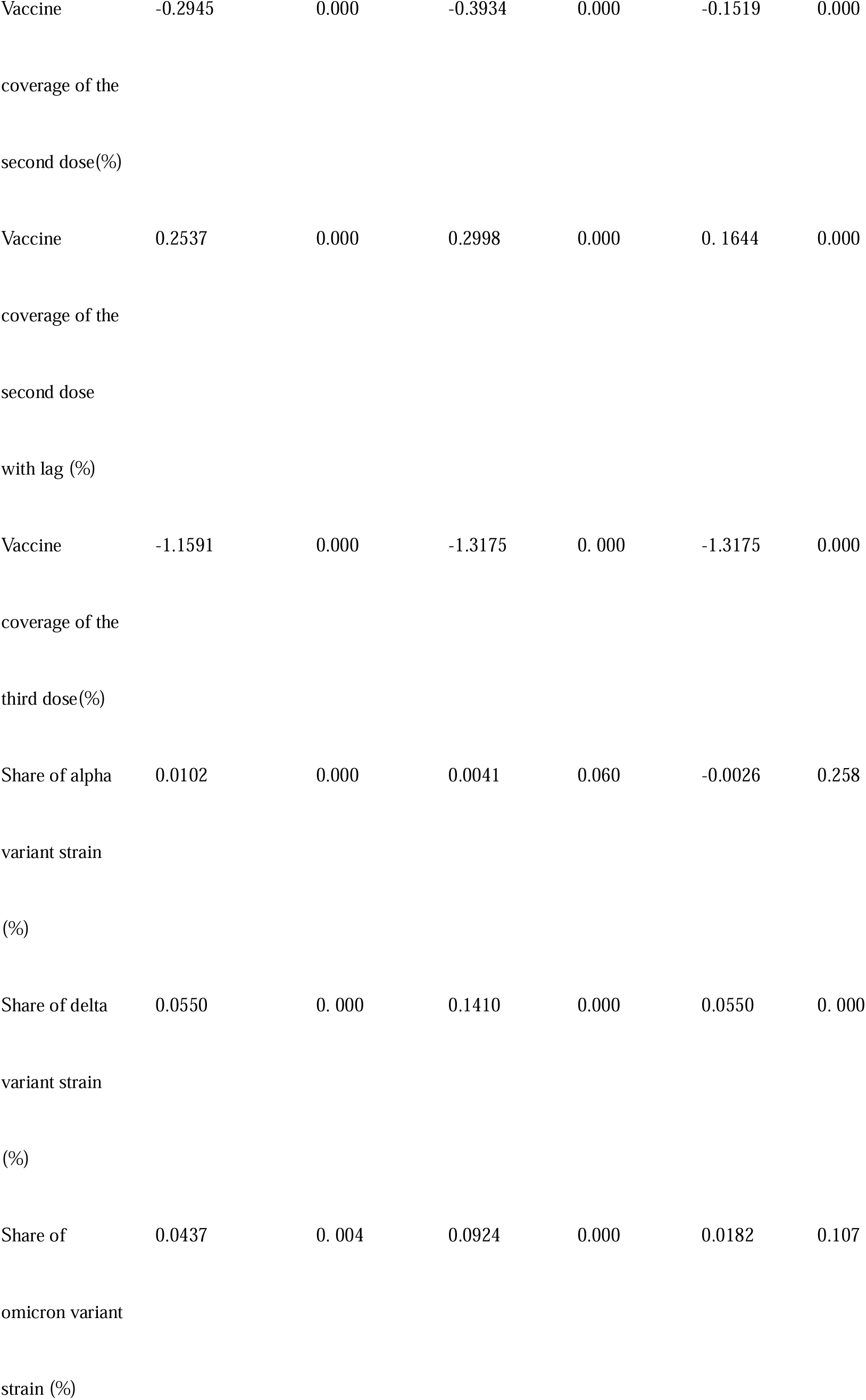
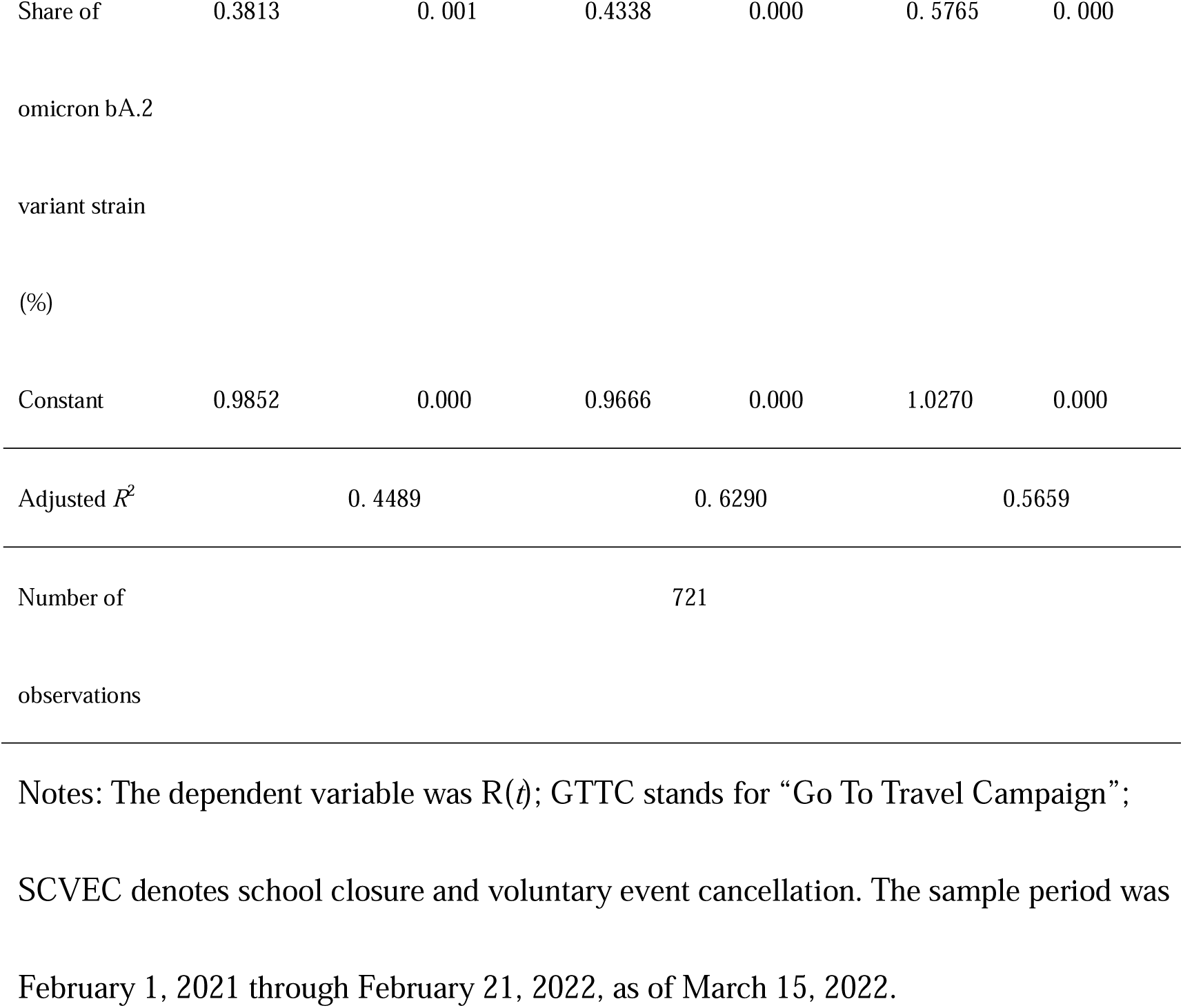
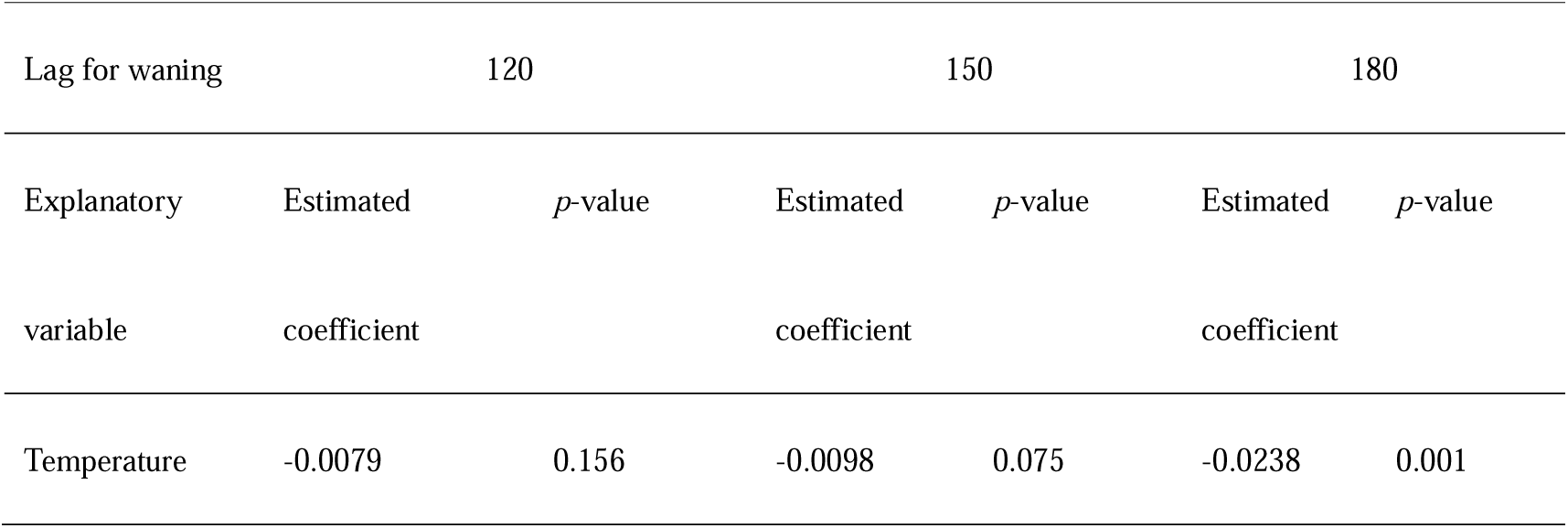
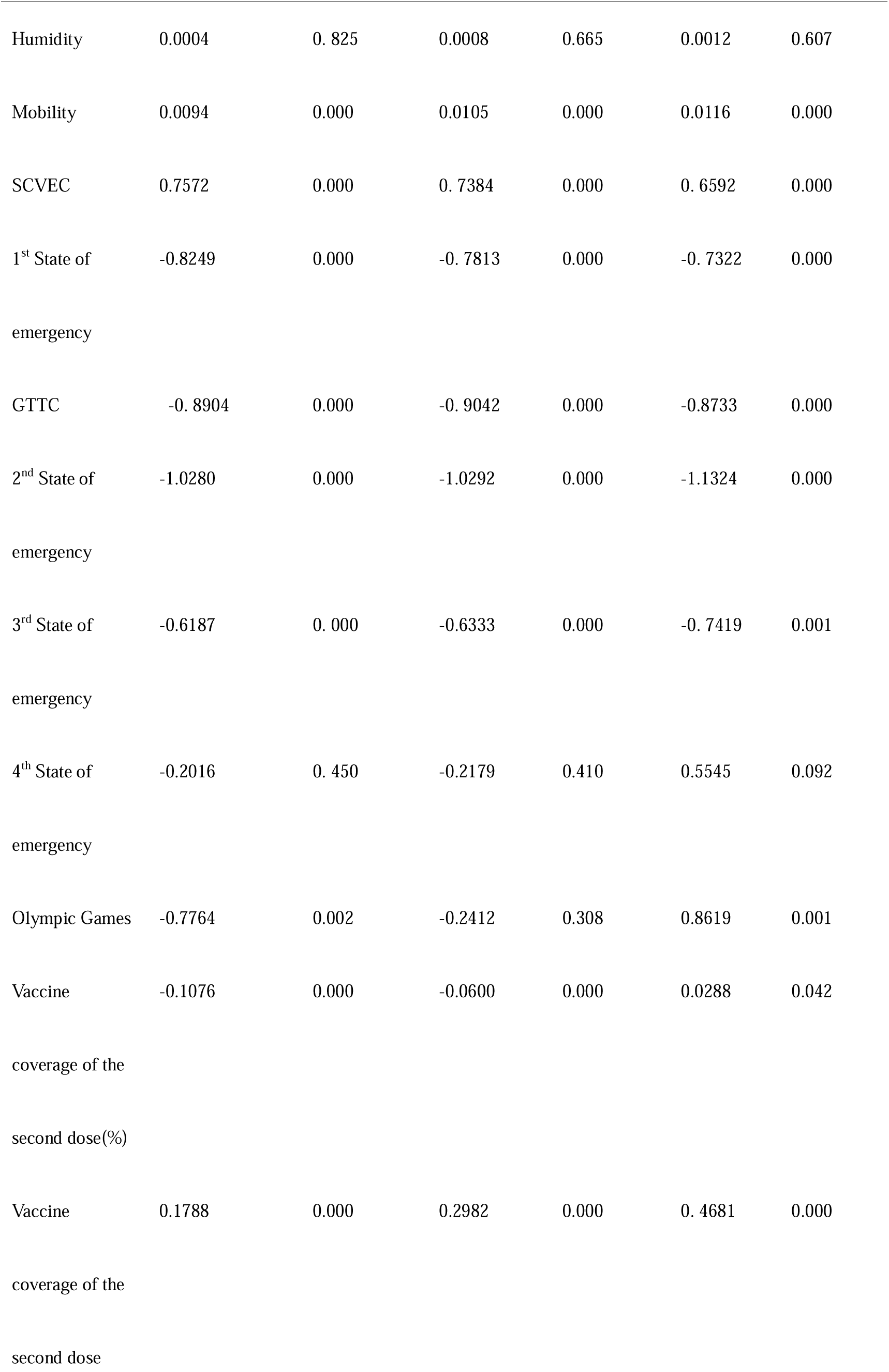
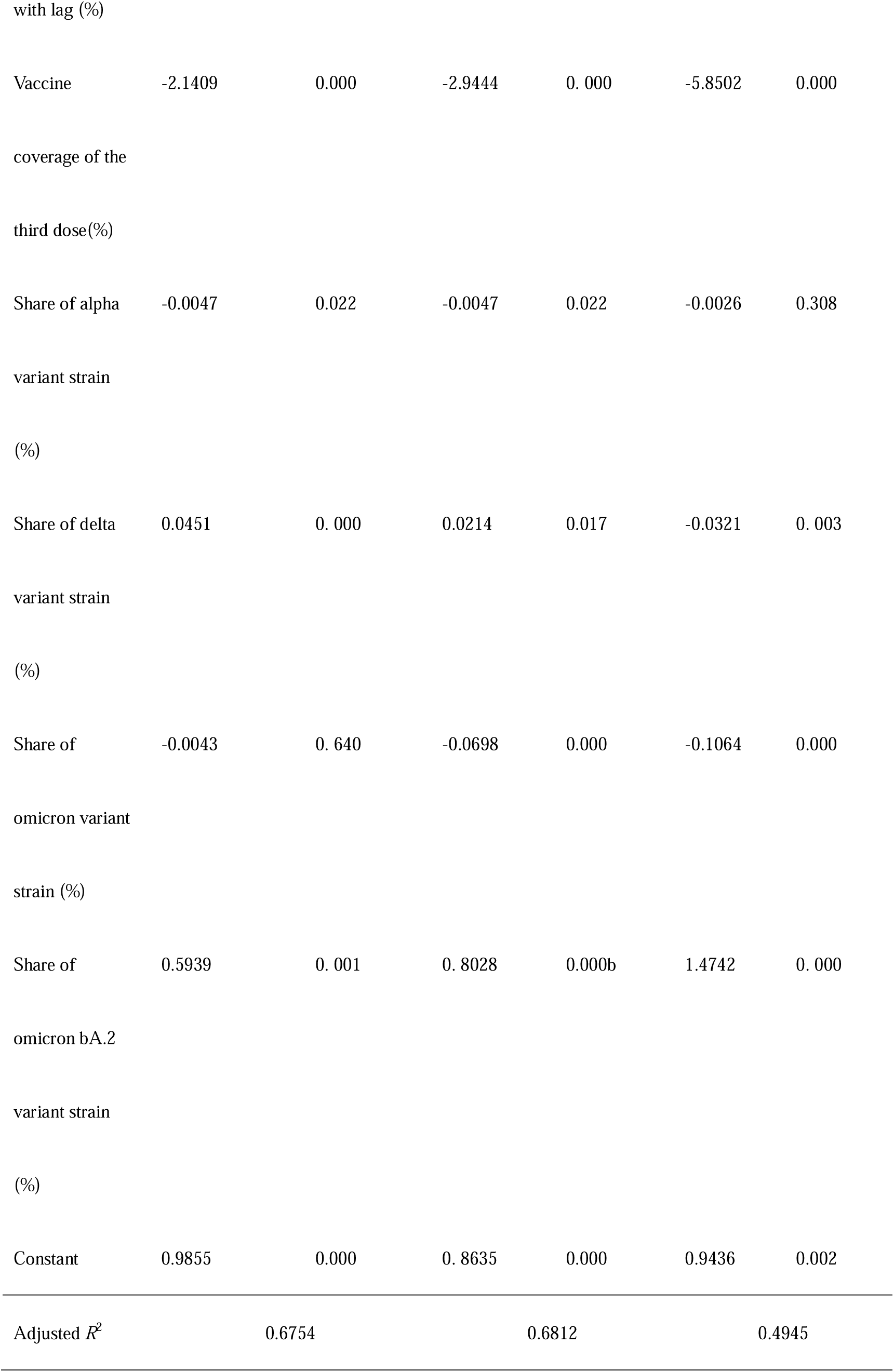
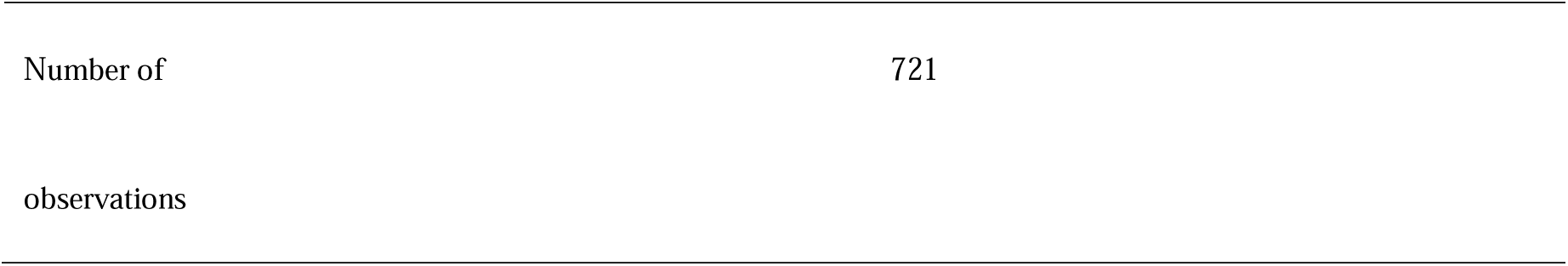
Estimation results of R(*t*) with vaccine coverage, prevalence of the variant strains, and Olympic Games with the climate condition, mobility, and countermeasures

| Lag for waning |  | 30 |  | 60 |  | 90 |
| --- | --- | --- | --- | --- | --- | --- |
| Explanatory | Estimated | *p*-value | Estimated | *p*-value | Estimated | *p*-value |
| variable | coefficient |  | coefficient |  | coefficient |  |
| Temperature | -0.00188 | 0.010 | -0.0086 | 0.148 | -0.0143 | 0.025 |
| Humidity | 0.0009 | 0.706 | 0.0013 | 0.522 | 0.0012 | 0.583 |
| Mobility | 0.0101 | 0.000 | 0.0086 | 0.000 | 0.0093 | 0.000 |
| SCVEC | 0.74542 | 0.000 | 0.8124 | 0.000 | 0.7363 | 0.000 |
| 1<sup>st</sup> State of emergency | -0.7331 | 0.000 | -0.8073 | 0.000 | -0.8155 | 0.000 |
| GTTC | -0.8090 | 0.000 | -0.8330 | 0.000 | -0.8630 | 0.000 |
| 2<sup>nd</sup> State of emergency | -1.0838 | 0.000 | -1.0020 | 0.000 | -1.0657 | 0.000 |
| 3<sup>rd</sup> State of emergency | -1.4772 | 0.000 | -1.0563 | 0.000 | -0.6957 | 0.000 |
| 4<sup>th</sup> State of emergency | 0.3742 | 0.282 | 0.8316 | 0.003 | 0.5655 | 0.063 |
| Olympic Games | 0.4432 | 0.159 | -1.9756 | 0.000 | -1.0490 | 0.000 |
| Vaccine | -0.2945 | 0.000 | -0.3934 | 0.000 | -0.1519 | 0.000 |
| coverage of the |  |  |  |  |  |  |
| second dose(%) |  |  |  |  |  |  |
| Vaccine | 0.2537 | 0.000 | 0.2998 | 0.000 | 0. 1644 | 0.000 |
| coverage of the |  |  |  |  |  |  |
| second dose |  |  |  |  |  |  |
| with lag (%) |  |  |  |  |  |  |
| Vaccine | -1.1591 | 0.000 | -1.3175 | 0. 000 | -1.3175 | 0.000 |
| coverage of the |  |  |  |  |  |  |
| third dose(%) |  |  |  |  |  |  |
| Share of alpha | 0.0102 | 0.000 | 0.0041 | 0.060 | -0.0026 | 0.258 |
| variant strain |  |  |  |  |  |  |
| (%) |  |  |  |  |  |  |
| Share of delta | 0.0550 | 0. 000 | 0.1410 | 0.000 | 0.0550 | 0. 000 |
| variant strain |  |  |  |  |  |  |
| (%) |  |  |  |  |  |  |
| Share of | 0.0437 | 0. 004 | 0.0924 | 0.000 | 0.0182 | 0.107 |
| omicron variant |  |  |  |  |  |  |
| strain (%) |  |  |  |  |  |  |
| Share of | 0.3813 | 0.001 | 0.4338 | 0.000 | 0.5765 | 0.000 |
| omicron bA.2 |  |  |  |  |  |  |
| variant strain |  |  |  |  |  |  |
| (%) |  |  |  |  |  |  |
| Constant | 0.9852 | 0.000 | 0.9666 | 0.000 | 1.0270 | 0.000 |
| Adjusted $R^2$ | | 0.4489 | | 0.6290 | | 0.5659 |
| Number of |  |  | 721 |  |  |  |
| observations |  |  |  |  |  |  |

Notes: The dependent variable was $R(t)$ ; GTTC stands for “Go To Travel Campaign”; SCVEC denotes school closure and voluntary event cancellation. The sample period was February 1, 2021 through February 21, 2022, as of March 15, 2022.

Table 1 (cont.)
|  |  |  |  |  |  |  |
| --- | --- | --- | --- | --- | --- | --- |
| Lag for waning | 120 |  | 150 |  | 180 |  |
| Explanatory | Estimated | $p$-value | Estimated | $p$-value | Estimated | $p$-value |
| variable | coefficient |  | coefficient |  | coefficient |  |
| Temperature | -0.0079 | 0.156 | -0.0098 | 0.075 | -0.0238 | 0.001 |
| Humidity | 0.0004 | 0. 825 | 0.0008 | 0.665 | 0.0012 | 0.607 |
| Mobility | 0.0094 | 0.000 | 0.0105 | 0.000 | 0.0116 | 0.000 |
| SCVEC | 0.7572 | 0.000 | 0. 7384 | 0.000 | 0. 6592 | 0.000 |
| 1<sup>st</sup> State of<br>emergency | -0.8249 | 0.000 | -0. 7813 | 0.000 | -0. 7322 | 0.000 |
| GTTC | -0. 8904 | 0.000 | -0. 9042 | 0.000 | -0.8733 | 0.000 |
| 2<sup>nd</sup> State of<br>emergency | -1.0280 | 0.000 | -1.0292 | 0.000 | -1.1324 | 0.000 |
| 3<sup>rd</sup> State of<br>emergency | -0.6187 | 0. 000 | -0.6333 | 0.000 | -0. 7419 | 0.001 |
| 4<sup>th</sup> State of<br>emergency | -0.2016 | 0. 450 | -0.2179 | 0.410 | 0.5545 | 0.092 |
| Olympic Games | -0.7764 | 0.002 | -0.2412 | 0.308 | 0.8619 | 0.001 |
| Vaccine<br>coverage of the<br>second dose(%) | -0.1076 | 0.000 | -0.0600 | 0.000 | 0.0288 | 0.042 |
| Vaccine<br>coverage of the<br>second dose | 0.1788 | 0.000 | 0.2982 | 0.000 | 0. 4681 | 0.000 |
| with lag (%) |  |  |  |  |  |  |
| Vaccine | -2.1409 | 0.000 | -2.9444 | 0.000 | -5.8502 | 0.000 |
| coverage of the |  |  |  |  |  |  |
| third dose(%) |  |  |  |  |  |  |
| Share of alpha | -0.0047 | 0.022 | -0.0047 | 0.022 | -0.0026 | 0.308 |
| variant strain |  |  |  |  |  |  |
| (%) |  |  |  |  |  |  |
| Share of delta | 0.0451 | 0.000 | 0.0214 | 0.017 | -0.0321 | 0.003 |
| variant strain |  |  |  |  |  |  |
| (%) |  |  |  |  |  |  |
| Share of | -0.0043 | 0.640 | -0.0698 | 0.000 | -0.1064 | 0.000 |
| omicron variant |  |  |  |  |  |  |
| strain (%) |  |  |  |  |  |  |
| Share of | 0.5939 | 0.001 | 0.8028 | 0.000b | 1.4742 | 0.000 |
| omicron bA.2 |  |  |  |  |  |  |
| variant strain |  |  |  |  |  |  |
| (%) |  |  |  |  |  |  |
| Constant | 0.9855 | 0.000 | 0.8635 | 0.000 | 0.9436 | 0.002 |
| Adjusted $R^2$ | | 0.6754 | | 0.6812 | | 0.4945 |

|  |  |
| --- | --- |
| Number of | 721 |
| observations |  |

Of significant coefficients, the estimated sign of mobility, share of delta or omicron bA.2 variant strain, the 1st to 3rd state of emergency, and vaccine coverage of the second or third dose were as expected, though the estimated sign of SCVEC, and share of alpha or omicron variant strain were unexpected.

## Discussion

The obtained estimated results showed that waning of the second dose vaccine e with 150 days prior was the most appropriate specification. This duration may be comparable with earlier studies of waning [8,9], which reached their conclusions based on antibody titer or test negative design. Readers must be reminded that waning estimated for the present study might include behavioral changes among the vaccinated persons to adoption of more risky behavior that is prone to exacerbating infectively. Such behaviors and the vaccine itself affect waning results, but they are not separately discernible based on results of this study. Weakening of immunoreaction and behavioral change are separate factors, but their mutual effects might be the most important for management of public health.

Moreover, the estimated infectively of bA.2 was quite high even though infectively of omicron was lower than traditional strain before alpha variant strain. This extremely result may be caused by data limited lower level of bA.2 prevalence due to the study period. In this sense, out obtained result might be interim results, and not conclusive. Hereafter, expanding of bA.2, its estimated infectively may be able to decrease over time. Vaccine efficacy was estimated as 95% for the original strain through clinical trials [30]. In the real world, it was also estimated as 46–80% for the first dose and 86–90% for the second dose [31-36] through case–control studies or test-negative design. However, even in the real world, such studies specifically examine protection for vaccine recipients only and ignore herd immunity, representing vaccine effects on non-vaccine recipients. The latter was not able to be estimated through clinical trials, case–control studies, or test negative design. In this sense, these earlier studies have been incapable of evaluating the overall effects of vaccination on the community. Instead of those methods, we evaluated vaccine effectiveness on the entire community, of course including herd immunity, through its effects on SARS-CoV-2 infectively.

Results indicated no significant result of momentary effects from the current vaccine coverage or waning from past vaccine coverage when the past vaccine coverage was defined as less than 60 days prior. Particularly, these estimated coefficients had unexpected signs but were significant. These results were probably caused by multicollinearity among the current and past vaccine coverage. Because of smaller time differences among these variables, correlation among them can be expected to be higher. Therefore, the smaller time differences distorted estimation results.

Conversely, when the past vaccine coverage was defined as more than 120 days prior, the estimated coefficients of the past vaccine coverages were much larger than the estimated coefficients of the current vaccine coverage in absolute terms. Statistically, the result probably reflected that the past vaccine coverage occurring longer ago should be a very small number, as shown in Figure 1. Therefore, the estimated coefficients should be larger than the correspondence when the past vaccine coverage was defined as 60 or 90 days prior.

Expressed semantically, because that waning might not reduce the immunization level to less than before the vaccination was administered, these results imply that behavioral changes to adopt more risky behaviors prone to infection among vaccinated persons raise infectively considerably. No evidence exists to indicate that the Tokyo Games 2020 exacerbated the outbreak of COVID-19. Expectations by some experts before the Olympic Games might have been wrong. It seems likely that most Japanese people watched TV at home and rooted for athletes. The no-audience policy might have contributed to reduction in infectively during the Games. Even though lower infectively prevailed during the Games, if it actually became higher than unity, then the number of newly infected or newly confirmed patients would be expected to grow during the period. Therefore, the number of patients has not represented the outbreak situation accurately. Infectively must be specifically examined during that period to evaluate policies adequately.

Alpha variant strain effects were significant and negative. Additionally, the share of delta variant strain was not found to be significant, with some exceptions. These results were not consistent with results reported from earlier studies [10-13].

### Limitations

First, we assumed implicitly that epidemiological characteristics including incubation period or delay in reports were the same among the original strain, alpha, delta, omicron and bA.2 variant strains. However, results of one study indicated that the delta variant strain has a shorter incubation period than either original strain [37].

Secondly, readers must be reminded when interpreting the obtained results that they do not indicate causality. Results of this study demonstrated that a negative association exists between the vaccine coverage and infectively. That finding does not necessarily mean that the vaccine coverage reduced infectively. The lower infectively might have caused or might have even simply coincided with higher vaccine coverage.

## Conclusion

We found that second dose vaccine coverage with 150 days prior raises infectively, and bA.2 of omicron variant strain has higher infectively than other variant strain or traditional strain. Because of data limitation since emerging bA.2, the estimated its infectively will change over time.

The present study is based on the authors’ opinions: it does not reflect any stance or policy of their professionally affiliated bodies.

## Data Availability

Japan Ministry of Health, Labour and Welfare. Press Releases.(in Japanese)

https://www.mhlw.go.jp/stf/newpage_10723.html

## Acknowledgments

We acknowledge the great efforts of all staff at public health centers, medical institutions, and other facilities fighting the spread and destruction associated with COVID-19.

## Ethical considerations

All information used for this study was from official data published on the internet. There is therefore no ethical issue related to this study.

## References

1. Araf Y, Akter F, Tang YD, Fatemi R, Parvez MSA, Zheng C, Hossain MG. Omicron variant of SARS-CoV-2: Genomics, transmissibility, and responses to current COVID-19 vaccines. J Med Virol 2022. 94:1825–32.

2. Ren SY, Wang WB, Gao RD, Zhou AM. Omicron variant (B.1.1.529) of SARS-CoV-2: Mutation, infectivity, transmission, and vaccine resistance. World J Clin Cases 2022.10:1–11. doi: 10.12998/wjcc.v10.i1.1.

3. Zhou H, Tada T, Dcosta BM, Landau NR. Neutralization of SARS-CoV-2 Omicron BA.2 by Therapeutic Monoclonal Antibodies. bioRxiv 2022.2022.02.15.480166. doi: 10.1101/2022.02.15.480166.

4. Cheng VC, Ip JD, Chu AW, Tam AR, Chan WM, Abdullah SMU, Chan BP, Wong SC, Kwan MY, Chua GT, Ip P, Chan JM, Lam BH, To WK, Chuang VW, Yuen KY, Hung IF, To KK. Rapid spread of SARS-CoV-2 Omicron subvariant BA.2 in a single-source community outbreak. Clin Infect Dis 2022.ciac203. doi: 10.1093/cid/ciac203.

5. Fonager J, Bennedbak M, Bager P, Wohlfahrt J, Ellegaard KM, Ingham AC, Edslev SM, Stegger M, Sieber RN, Lassauniere R, Fomsgaard A, Lillebaek T, Svarrer CW, Moller FT, Moller CH, Legarth R, Sydenham TV, Steinke K, Paulsen SJ, Castruita JAS, Schneider UV, Schouw CH, Nielsen XC, Overvad M, Nielsen RT, Marvig RL, Pedersen MS, Nielsen L, Nilsson LL, Bybjerg-Grauholm J, Tarpgaard IH, Ebsen TS, Lam JUH, Gunalan V, Rasmussen M. Molecular epidemiology of the SARS-CoV-2 variant Omicron BA.2 sub-lineage in Denmark, 29 November 2021 to 2 January 2022. Euro Surveill 2022.27:2200181. doi: 10.2807/1560-7917.ES.2022.27.10.2200181.

6. Prime Minister and his Cabinet. Novel Coronavirus Vaccines. https://www.kantei.go.jp/jp/headline/kansensho/vaccine.html (in Japanese) (accessed Nov 30, 2021)

7. Japan Ministry of Health, Labour and Welfare. Situation of vaccine coverage for COVID-19. https://www.mhlw.go.jp/stf/seisakunitsuite/bunya/vaccine_sesshujisseki.html (in Japanese) (accessed Nov 30, 2021)

8. Levin EG, Lustig Y, Cohen C, et al. Waning Immune Humoral Response to BNT162b2 Covid-19 Vaccine over 6 Months. N Engl J Med 2021:NEJMoa2114583.

9. Chemaitelly H, Tang P, Hasan MR, et al. Waning of BNT162b2 Vaccine Protection against SARS-CoV-2 Infection in Qatar. N Engl J Med. 2021:NEJMoa2114114.

10. Leung K, Shum MHH, Leung GM, Lam TTY, Wu JT. Early transmissibility assessment of the alpha variant strain of SARS-CoV-2 in the United Kingdom, October to November 2020. Euro Surveill 2021;26:2002106.

11. Graham MS, Sudre CH, May A, et al. Changes in symptomatology, reinfection, and transmissibility associated with the SARS-CoV-2 variant B.1.1.7: an ecological study. Lancet Public Health 2021;6:e335–e345.

12. Davies NG, Abbott S, Barnard RC, et al. Estimated transmissibility and impact of SARS-CoV-2 lineage B.1.1.7 in England. Science 2021;372(6538): eabg3055.

13. Zhao S, Lou J, Cao L, Zheng H, Chong MKC, Chen Z, Chan RWY, Zee BCY, Chan PKS, Wang MH. Quantifying the transmission advantage associated with alpha variant strain substitution of SARS-CoV-2 in the UK: an early data-driven analysis. J Travel Med 2021;28:taab011.

14. Hoang VT, Al-Tawfiq JA, Gautret P. The Tokyo Olympic Games and the Risk of COVID-19. Curr Trop Med Rep 2020;1–7.

15. Anzai A. “Go To Travel” Campaign and Travel-Associated Coronavirus Disease 2019 Cases: A Descriptive Analysis, July–August 2020. J. Clin. Med 2021;10:398. https://doi.org/10.3390/jcm10030398

16. Shi P, Dong Y, Yan H, Zhao C, Li X, Liu W, He M, Tang S, Xi S. Impact of temperature on the dynamics of the COVID-19 outbreak in China. Sci Total Environ. 2020;728:138890.

17. Tobias A, Molina T. Is temperature reducing the transmission of COVID-19? Environ Res. 2020;186:109553.

18. Yao Y, Pan J, Liu Z, Meng X, Wang W, Kan H, Wang W. No association of COVID-19 transmission with temperature or UV radiation in Chinese cities. Eur Respir J. 2020;55:2000517.

19. Walrand S. Autumn COVID-19 surge dates in Europe correlated to latitudes, not to temperature-humidity, pointing to vitamin D as contributing factor. Scientific Reports 2021;11:1981.

20. Kurita J, Sugishita Y, Sugawara T, Ohkusa Y. Mobility data can reveal the entire COVID1-19 outbreak course in Japan.JMIR Public Health & Surveillance 2021;7. https://publichealth.jmir.org/2021/2/e20335

21. Bergman N, Fishman R. Mobility Reduction and Covid-19 Transmission Rates. https://www.medrxiv.org/content/10.1101/2020.05.06.20093039v3

22. Flaxman S, Mishra S, Gandy A, Unwin HJT, Mellan TA, Coupland H, Whittaker C, Zhu H, Berah T, Eaton JW, Monod M, Imperial College COVID-19 Response Team; Ghani AC, Donnelly CA, Riley S, et al. Estimating the effects of non-pharmaceutical interventions on COVID-19 in Europe. Nature 2020;584:257–61.

23. Li Y, Campbell H, Kulkarni D, Harpur A, Nundy M, Wang X, Nair H, for theUsher Network for COVID-19 Evidence Reviews (UNCOVER) group. The temporal association of introducing and lifting non-pharmaceutical interventions with the time-varying reproduction number (R) of SARS-CoV-2: a modelling study across 131 countries. Lancet Infect Dis 2021;21:193–202

24. Larrosa JMC. SARS-CoV-2 in Argentina: Lockdown, mobility, and contagion. J Med Virol 2020.;93:2252–61.

25. Japan Ministry of Health, Labour and Welfare. Press Releases. https://www.mhlw.go.jp/stf/newpage_10723.html (in Japanese) (accessed Jan18, 2022).

26. Kurita J, Sugawara T, Ohkusa Y. Estimated effectiveness of school closure and voluntary event cancellation as COVID-19 countermeasures in Japan. J Infect Chemother 2021;27:62–4. doi: 10.1016/j.jiac.2020.08.012.

27. Sugishita Y, Kurita J, Sugawara T, Ohkusa Y. Effects of voluntary event cancellation and school closure as countermeasures against COVID-19 outbreak in Japan. PLOS One 2020.

28. Kimball A, Hatfield KM, Arons M, et al. Asymptomatic and Presymptomatic SARS-CoV-2 Infections in Residents of a Long-Term Care Skilled Nursing Facility – King County, Washington, March 2020. Morb Mortal Wkly Rep 2020;69:377–81.

29. Tokyo metropolitan Government. Data of COVID-19 monitoring meeting in metropolitan Tokyo. https://www.bousai.metro.tokyo.lg.jp/taisaku/saigai/1013388/index.html (in Japanese) (accessed April 21,2022)

30. Polack FP, Thomas SJ, Kitchin N, et al. Safety and Efficacy of the BNT162b2 mRNA Covid-19 Vaccine. N Engl J Med. 2020;383:2603–15.

31. Chung H, He S, Nasreen S, et al. Effectiveness of BNT162b2 and mRNA-1273 covid-19 vaccines against symptomatic SARS-CoV-2 infection and severe covid-19 outcomes in Ontario, Canada: test negative design study. BMJ 2021;374.

32. Dagan N, Barda N, Kepten E, Miron O, Perchik S, Katz MA, Hernan MA, Lipsitch M, Reis B, Balicer RD. BNT162b2 mRNA Covid-19 vaccine in a nationwide mass vaccination setting. N Engl J Med 2021;384:1412–23.

33. Vasileiou E, Simpson CR, Shi T, et al. Interim findings from first-dose mass COVID-19 vaccination roll-out and COVID-19 hospital admissions in Scotland: a national prospective cohort study. Lancet 2021;397:1646–57.

34. Bernal JL, Andrews N, Gower C, Robertson C, Stowe J, Tessier E, Simmons R, Cottrell S, Roberts R, O’Doherty M, Brown K, Cameron C, Stockton D, McMenamin J, Ramsay M. Effectiveness of the Pfizer–BioNTech and Oxford–AstraZeneca vaccines on Covid-19 related symptoms, hospital admissions, and mortality in older adults in England: test negative case-control study. BMJ 2021;373:n1088.

35. Bjork J, Inghammar M, Moghaddassi M, Rasmussen M, Malmqvist U, Kahn F. Effectiveness of the BNT162b2 vaccine in preventing COVID-19 in the working age population: first results from a cohort study in southern Sweden. Infect Dis (Lond) 2021;1-6.

36. Pawlowski C, Lenehan P, Puranik A, Agarwal V, Venkatakrishnan AJ, Niesen MJM, O’Horo JC, Virk A, Swift MD, Badley AD, Halamka J, Soundararajan V. FDA-authorized COVID-19 vaccines are effective per real-world evidence synthesized across a multi-state health system. Med (N Y) 2021;2:979-92.e8.

37. Li B, Deng A, Li K, et al. Viral infection and transmission in a large, well-traced outbreak caused by the SARS-CoV-2 Delta variant. medRxiv 2021.07.07.21260122; doi: https://doi.org/10.1101/2021.07.07.21260122

